# Ischemia modulation via coronary revascularization and effects on the arrhythmic substrate

**DOI:** 10.1101/2025.04.17.25326044

**Authors:** Holly Morgan, Amedeo Chiribiri, Marina Strocchi, Hassan Zaidi, Nathan C. K. Wong, Azizah Ardinal, Mark Elliott, Steven Niederer, Matthew Ryan, Martin Bishop, Christopher Aldo Rinaldi, Divaka Perera

## Abstract

**Background:** Both ischemia and scar can contribute to arrhythmogenesis in patients with ischemic left ventricular dysfunction (ILVD); coronary revascularization is frequently undertaken to modify the former. Whether modulation of ischemia affects the substrate for ventricular arrhythmia is unclear.

**Objectives:** To assess the mechanistic effects of ischemia modulation via revascularization on the arrhythmic substrate in patients with ILVD.

**Methods:** Patients were eligible for enrolment if they had a left ventricular ejection fraction (LVEF)≤40%, extensive coronary disease (BCIS jeopardy score >6/12) and were scheduled to undergo percutaneous coronary intervention (PCI) or coronary artery bypass surgery (CABG). Scar and ischemic burden were assessed via stress-perfusion cardiac magnetic resonance (pCMR), calculated as a percentage of total LV myocardial volume. Arrhythmic substrate was characterised by non-invasive electrocardiographic imaging (ECGi). ECGi and pCMR were repeated 3 months after revascularization. The primary outcome was change in the LV activation recovery interval (ARI) dispersion.

**Results:** Thirty patients were enrolled (age 67±10 years, 87% male, LVEF 29±7%); 12 (40%) underwent CABG, 18 (60%) had PCI. Following revascularization, LVEF increased (+8±8%) and ischemic burden reduced (-34±24%)(p<0.01). There was no change in mean LV ARI dispersion, however individual changes in LV ARI dispersion correlated with individual changes in ischemic burden (r=0.51,p<0.01). Baseline LV volumes, scar burden, delta LVESVi and delta ischemia were all predictors of improvement.

**Conclusions:** Arrhythmic substrate was correlated with scar burden and was not altered by revascularization. Revascularization may not routinely reduce arrhythmic risk. Further work is needed to prospectively identify select patients who may confer benefit.

## Introduction

The primary cause of death in the ischemic left ventricular dysfunction (ILVD) population is sudden death related to ventricular arrhythmias. Both ischemia and scar can contribute to arrhythmogenesis but as the former is the only modifiable factor at present, ischemia modulation is often a focus for treatment. It has long been assumed that the burden of ischemia is directly correlated with clinical outcomes, and that treatment of this will reverse hibernation and reduce mortality.^1^ This is partly an extrapolation from the observation that successful primary percutaneous coronary intervention (PCI) in the setting of acute coronary syndromes reduces the risk of fatal and non-fatal ventricular arrhythmias. Therefore, despite absence of randomised trial data, revascularization is frequently undertaken in stable ILVD patients to reduce ventricular arrhythmia (VA) risk, and is ensconced in contemporary heart failure guidelines, as a class 1 level B recommendation.^2,3^

Observational studies of ILVD patients have reported lower mortality and arrhythmia rates in those who underwent revascularization prior to ICD insertion^4,5^; however such data are likely to be significantly confounded.^6^ In the Surgical Treatment for Ischemic Heart Failure (STICH) trial the occurrence of sudden death was lower in patients receiving coronary artery bypass grafting (CABG), with the benefit particularly apparent in patients surviving more than two years after randomisation.^7,8^ Notably the trial had a low rate of implantable cardioverter defibrillator (ICD) implantation and therefore cannot provide detailed evidence regarding VA occurrence or it’s alteration by revascularization. The Revascularization for Ischemic Ventricular Dysfunction – British Cardiovascular Intervention Society (REVIVED-BCIS2) trial found that in patients with ILVD, PCI did not reduce the occurrence of death or arrhythmic events over medical therapy alone.^9^ In small studies, alterations in the electrophysiological properties of the myocardium after revascularization have been found to occur earlier than, or independent from, left ventricular function recovery.^10^ In a sub-analysis of the EXPLORE trial, QT dispersion was reduced 4 months following chronic total occlusion PCI in patients with ILVD, independent of changes in left ventricular ejection fraction (LVEF).^11^

This study aimed to assess whether ischemia modulation via revascularization alters the arrhythmic substrate in patients with ILVD.

## Methods

### Study design

Patients with extensive coronary disease (defined by a BCIS jeopardy score of ≥6 out of 12) and LVEF ≤ 40% who were due to undergo revascularization with either CABG or PCI were enrolled. Exclusion criteria included acute myocardial infarction or haemodynamic instability in the preceding 72 hours, pregnancy or contraindication for cardiac magnetic resonance (CMR).

The study protocol was approved by the UK Health Research Authority in 2021; research ethics committee approval was gained prior to study commencement (REC reference: 21/SC/0024; IRAS 281825). All participants provided written informed consent. At the first study visit, demographic data, baseline New York Heart Association (NYHA) and Canadian Cardiovascular Society (CCS) class and 12 lead ECG were recorded. Patients then underwent electrocardiographic imaging (ECGi) and stress perfusion CMR. Revascularization was subsequently undertaken as per clinical indication with either PCI or CABG. Three months post revascularization, patients returned for repeat ECGi and CMR. The revascularization index was calculated for both modalities as (baseline BCIS-JS – post revascularization BCIS-JS) / baseline BCIS-JS *100.^12,13^

### Electrocardiographic imaging and measurement of arrhythmic substrate

Body surface potentials were recorded using the 252-electrode CardioInsight vest (Medtronic, Minnesota, USA), approved for research use in the UK and commercial use in the United States.^14^ Anatomical co-registration was then undertaken via non-contrast computed tomography thorax and the epicardial potentials reconstructed from body surface potentials and heart-torso geometry. Epicardial maps were analysed with Python3 and custom developed code used to filter electrograms and calculate activation time (AT), repolarisation time (RT) and activation recovery interval (ARI; a surrogate of local action-potential duration) at each ventricular epicardial point; this methodology has been previously described by our group.^15^ Readers were blinded to clinical or CMR data, revascularization modality or the temporal sequence of the ECGi study (ie baseline or follow up). Repolarisation times and ARIs were corrected for heart rate using the Fridericia formula.^16^ Biventricular (global) as well as LV specific ARI were computed as the difference between the corrected local repolarisation and activation times (Table S1). Biventricular and LV ARI dispersion were computed as the standard deviation of the ARIs.

### CMR protocol and analysis

CMR was preferentially performed on a 3-Tesla scanner (Achieva, Philips Healthcare, Netherlands) to provide high resolution, reduced signal-to-noise and consistency for analysis. Stress perfusion data were acquired using dual sequence implementation with ECG triggering. After establishing resting heart rate and blood pressure, adenosine was infused at a rate of 140 μg/kg/min for at least three minutes or until adequate hyperaemia was achieved, with dose escalation to 175 then 210 μg/kg/min if required. High-resolution images were acquired in three short-axis slices covering the left ventricle in addition to the low-resolution AIF slice. A minimum of 10 minutes after the stress scan, rest sequences were taken before acquisition of late gadolinium enhancement (LGE) images; both bright and dark blood sequences were performed. LV end-systolic volume (LVESV) and LV end-diastolic volume (LVEDV) were indexed to body surface area (Mosteller formula). Scar and ischemic burden were visually assessed per AHA segment and are presented as a percentage of total LV myocardial volume (Table S2). Readers were blinded to clinical or ECGi data, revascularization modality or the temporal sequence of the CMR study.

### Outcomes

The primary outcome measure was the change in arrhythmic substrate, from baseline to post revascularization, referred to as the delta LV ARI dispersion. Secondary outcome measures included delta LV ARI, delta ARI, delta biventricular ARI dispersion, delta LVEF and delta LV end systolic volume.

Participants were split into two groups: ‘improved’ if delta LV ARI dispersion was less than 0, and ‘failed to improve’ if delta LV ARI dispersion was greater than or equal to 0.

### Statistical methods

A power calculation was undertaken using delta LV ARI as the dependent variable and delta ischemic burden as the independent variable. This identified that a sample size of 29 would provide 80% power (at 5% significance) to demonstrate a correlation between delta LV ARI and delta ischemic burden, where rho is not smaller than 0.5 (with a 95% confidence interval of 0.16 to 0.74).^17^

Categorical demographic data are presented as counts (percentages) and continuous data as means (standard deviations) or medians [interquartile ranges] depending on the normality of distribution. Categorical variables were compared using the chi-squared test. Continuous paired endpoints were compared using paired (within groups) t-tests. When comparing sub-group delta values, unpaired (between groups) t-tests were used and findings reported as the difference in mean change. Correlations between metrics were assessed using Spearman’s correlation coefficient. Binary logistic regression, variable importance models and multiple logistic regression were used to assess variables associated with functional recovery. All analyses were conducted using SPSS, GraphPad Prism and R studio. A p-value of <0.05 was considered significant.

## Results

### Baseline characteristics

Between March 2021 and November 2023, 30 patients were enrolled (Figure 1); participant demographics are shown in Table 1. At baseline, participants had severely impaired LVEF (29 ± 7%), with a high burden of scar (17 ± 11%) and ischemia (50 ± 19%)(Table 2). Twelve participants underwent CABG (40%) and 18 had PCI (60%). Participants undergoing CABG and PCI had similar characteristics, except for LVEF which was significantly higher in patients undergoing CABG (Table 1).

**Figure 1.**
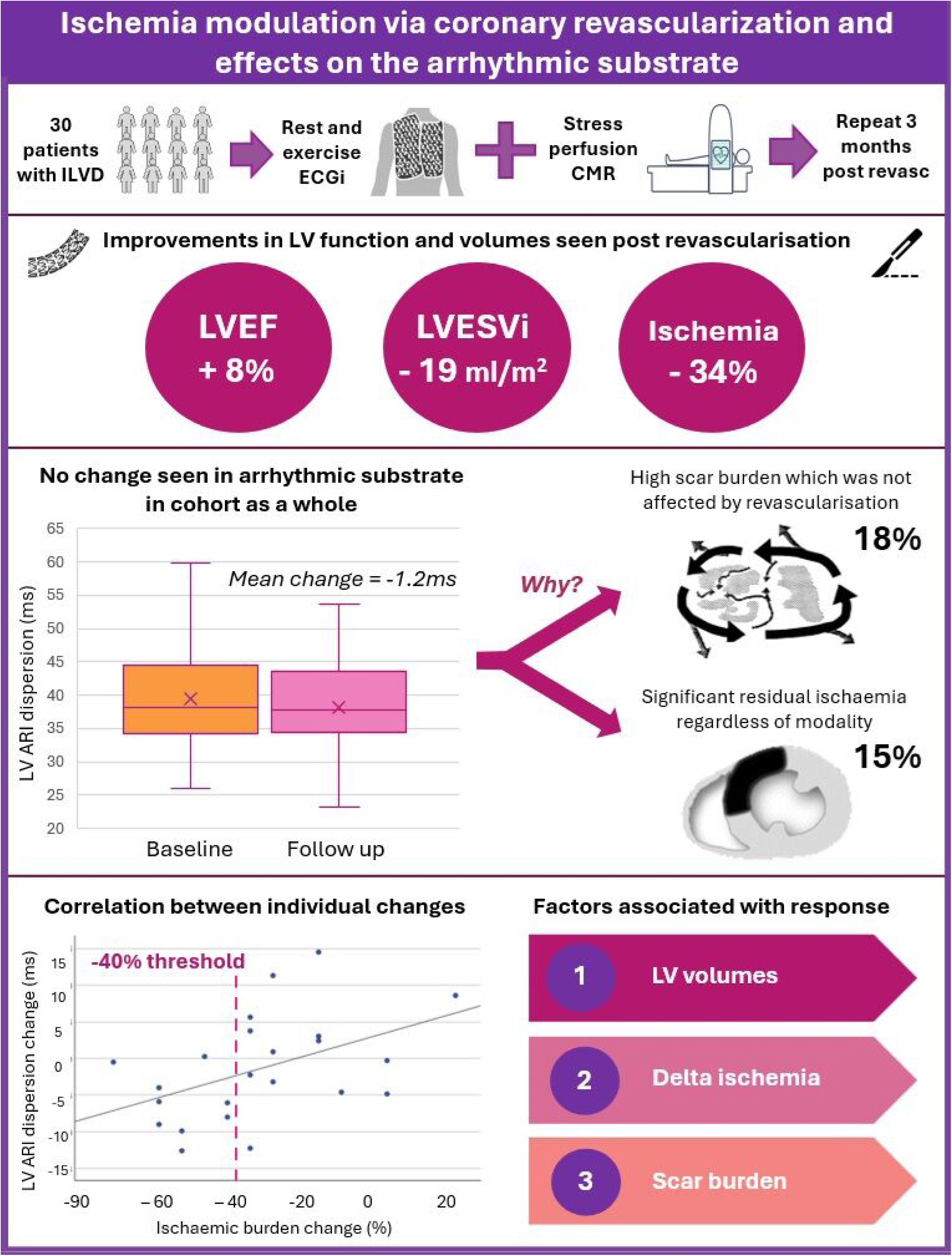
Study CONSORT.

**Table 1.** Patient demographics, divided by revascularization modality.

|  | All (n=30) | PCI (n=18) | CABG (n=12) | P value |
| --- | --- | --- | --- | --- |
| **Age – yrs** | 66.5 ± 10.1 | 66.9 ± 9.8 | 65.9 ± 10.6 | 0.62 |
| **Male sex – no (%)** | 26 (87) | 16 (89) | 10 (83) | 0.81 |
| **NYHA functional class – no (%)** |  |  |  | 0.32 |
| I or II | 19 (63) | 11 (61) | 8 (67) |  |
| III or IV | 11 (37) | 7 (39) | 4 (33) |  |
| **CCS angina class – no (%)** |  |  |  | 0.76 |
| I or II | 13 (43) | 6 (33) | 7 (58) |  |
| III or IV | 17 (57) | 12 (67) | 5 (42) |  |
| **Body mass index** | 28.4 ± 4.6 | 27.4 ± 3.5 | 29.9 ± 5.6 | 0.16 |
| **Hypertension – no (%)** | 21 (70) | 12 (67) | 9 (75) | 0.64 |
| **Diabetes – no (%)** | 14 (47) | 7 (39) | 7 (58) | 0.30 |
| **Previous revascularization – no (%)\*** | 5 (17) | 3 (17) | 2 (17) | 1.00 |
| **LVEF on echocardiography** | 29 ± 9 | 26.4 ± 8.0 | 32.5 ± 8.0 | 0.06* |
| **BCIS jeopardy score<sup>†</sup> – [IQR]** | 10 [8-12] | 10 [8-12] | 10 [10-12] | 0.83 |
| **Beta blockers – no (%)** | 24 (80) | 14 (78) | 10 (83) | 0.71 |
| **ACE/ARB/ARNI – no (%)** | 28 (93) | 17 (94) | 11 (92) | 0.77 |
| **MRA – no (%)** | 18 (60) | 12 (67) | 6 (50) | 0.36 |
| **SGLT2 inhibitors – no (%)** | 17 (57) | 12 (67) | 5 (42) | 0.18 |
| **QRS duration (ms)** | 111 ± 23 | 107 ± 25 | 114 ± 16 | 0.40 |
| **ACS indication for revasc – no (%)** | 12 (40) | 7 (39) | 5 (42) | 0.88 |

NYHA: New York Heart Association, CCS: Canadian Cardiovascular Society, BCIS: British Cardiovascular Intervention Society, LVEF: Left Ventricular Ejection Fraction, ACE: Angiotensin Converting Enzyme inhibitor, ARB: Angiotensin Receptor Blocker, ARNI: Angiotensin Receptor/Neprilysin Inhibitors, MRA: Mineralocorticoid Receptor Antagonist, SGLT2: Sodium Glucose Transport Protein 2 inhibitors, ACS: Acute Coronary Syndrome
\*All previous revascularisation was via PCI. <sup>†</sup>BCIS jeopardy score maximum possible score is 12

**Table 2.**
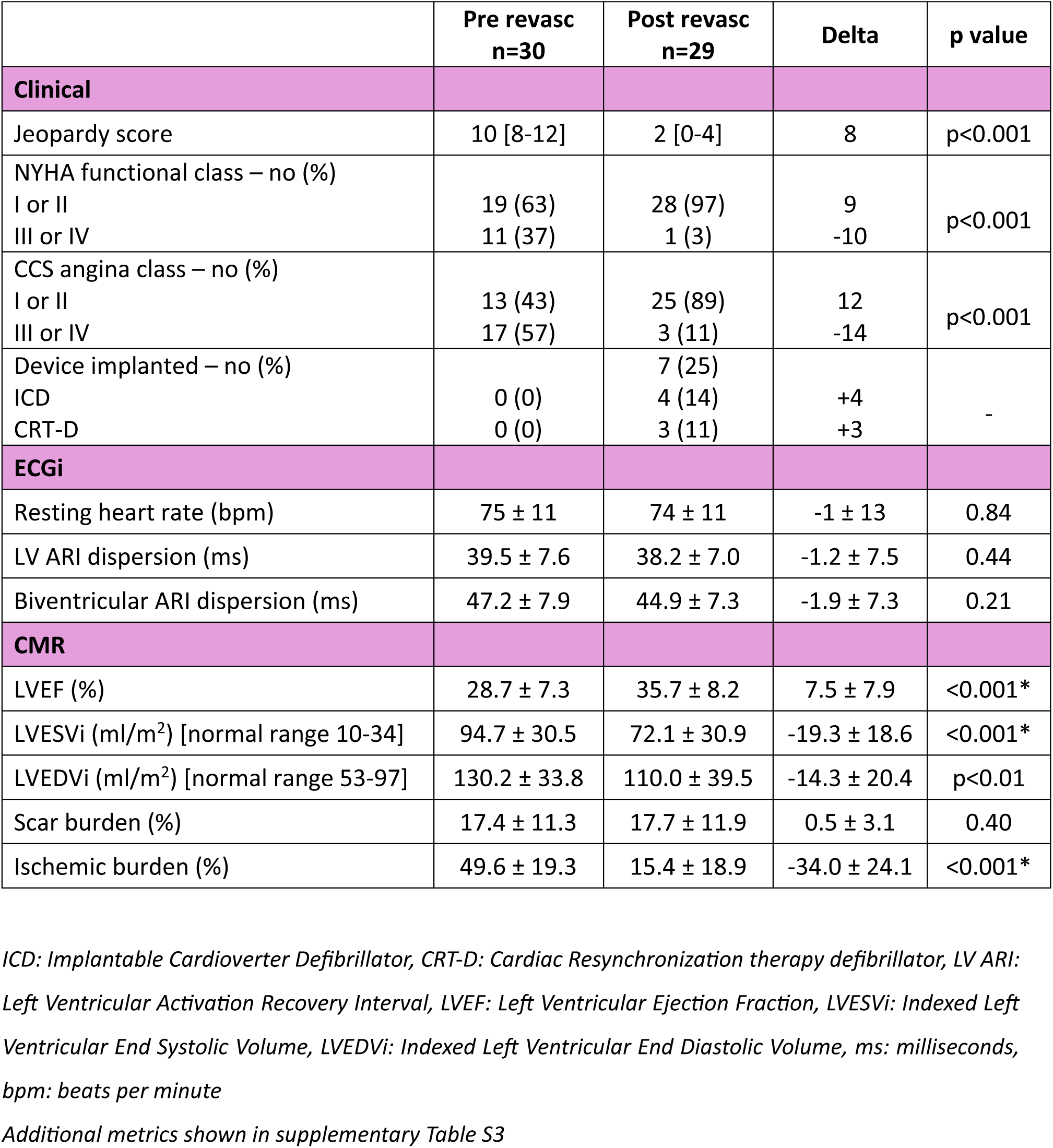
Baseline and follow up clinical, ECGi and CMR data.

ECGi data at baseline are summarised in Table 2. ECG QRS duration was correlated with LV total activation time on ECGi (r= 0.63, p<0.01, Figure S1). LV ARI dispersion was 40 ± 8ms and did not correlate with LVEF (r= -0.29, p=0.13) or ischemic burden (r= 0.28, p=0.17) but did correlate with scar burden (r=0.36, p=0.05)(Figure S2). Each 10% increase in scar burden was associated with an increase in LV ARI dispersion of 2.3ms (CI: 0.26 to 4.86).

### Impact of revascularization

Post-revascularization follow up was completed in 29 patients (97%); one patient died prior to follow up. Time from revascularization to follow up assessment was 128 days [106–138]. The coronary revascularization index was higher in those who underwent CABG (83 ± 17% versus 62 ± 18.4% for PCI, p<0.01). None of the patients had an implanted cardiac device at enrolment, however 25% had a device implanted during the follow-up period.

Patients reported improvement in cardiac symptoms, with reductions in NYHA and CCS angina class scores (Table 2). There was an improvement of 8% in LVEF, decrease of 19ml in LVESVi and 14ml in LVEDVi (Figure 2). Scar burden and scar entropy were unchanged, however ischemic burden reduced by -34 ± 24% (Table 2, Table S3).

**Figure 2.**
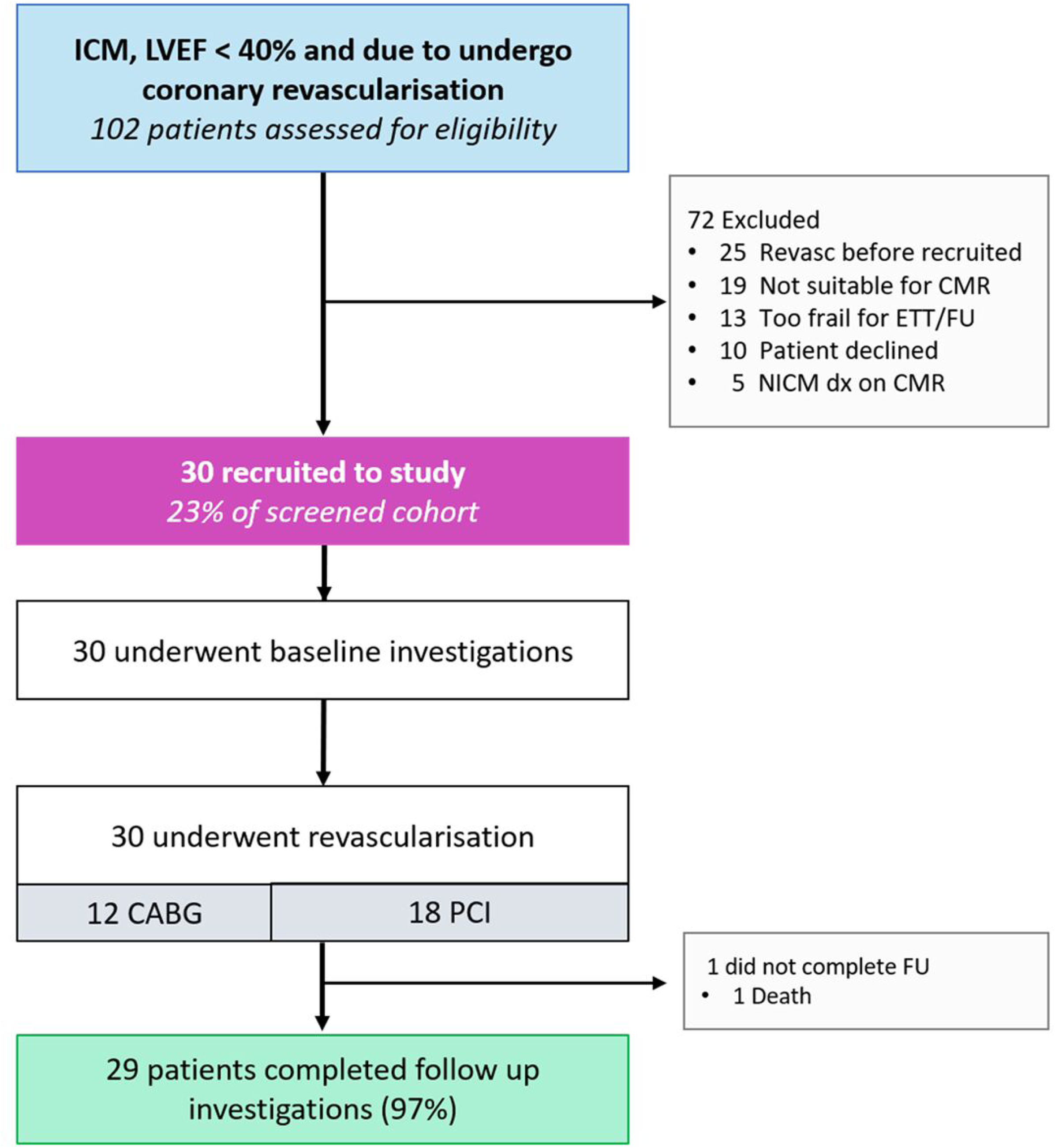
Change in LVEF, LVEDVi and LVESVi from pre to post revascularization.

A greater reduction in LV volumes were seen in patients who had undergone revascularization due to an acute coronary syndrome (LVESVi -30 ± 16 vs 12 ± 17, p<0.01)(Table S4). The change in ECGi and CMR metrics was similar in those who had undergone CABG versus PCI (Table S5).

### Primary and secondary outcomes

There was no overall change after revascularization in the mean LV ARI dispersion, LV ARI, biventricular ARI or biventricular ARI dispersion when considering the cohort as a whole (Figure 3, Table 2). However, changes in LV ARI dispersion were heterogeneous and individual change in ischemic burden was correlated with individual change in LV ARI dispersion (r=0.51, p<0.01)(Figure 4).

**Figure 3.**
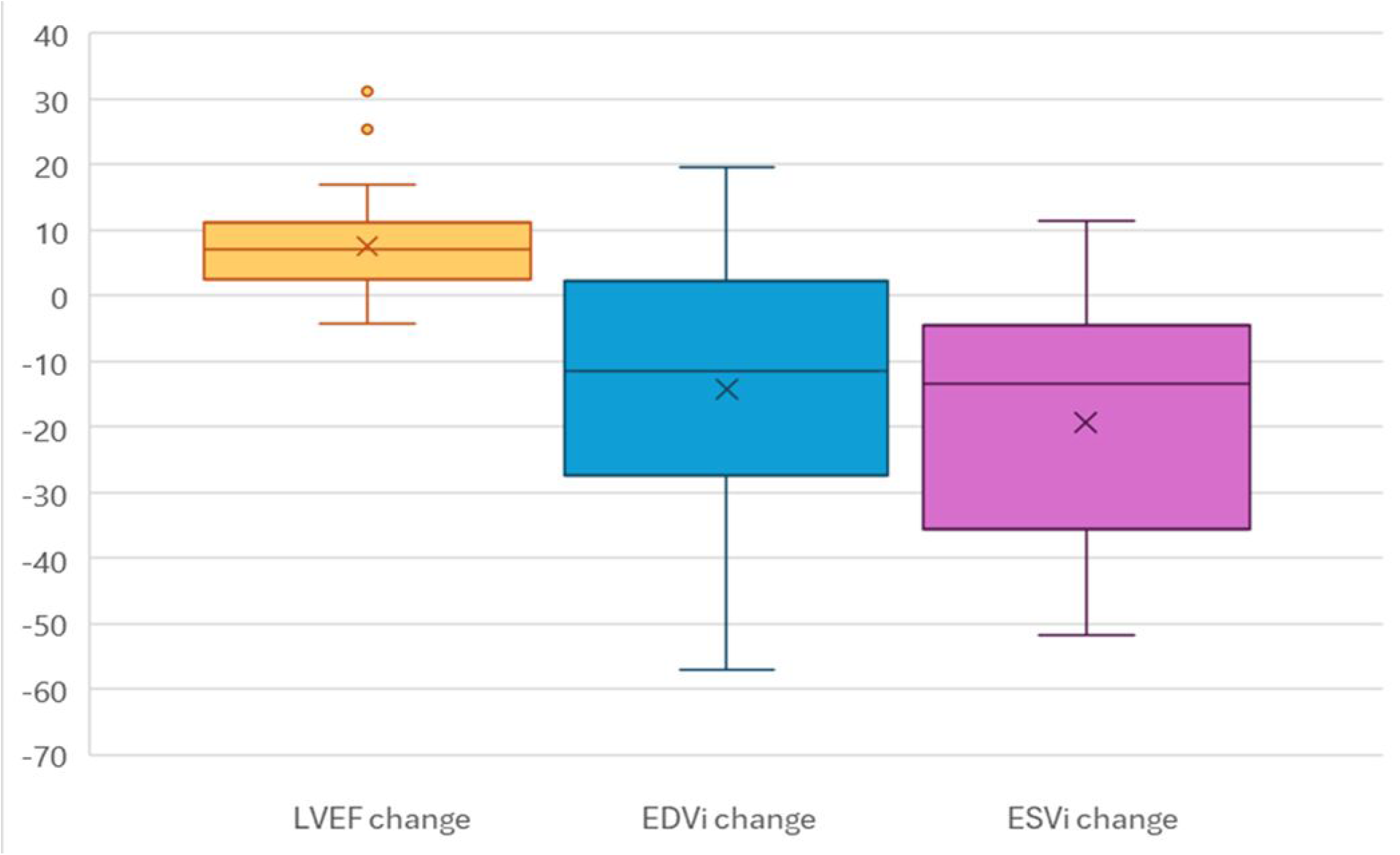
Change in biventricular ARI dispersion and LV ARI dispersion, pre and post revascularization.

**Figure 4.**
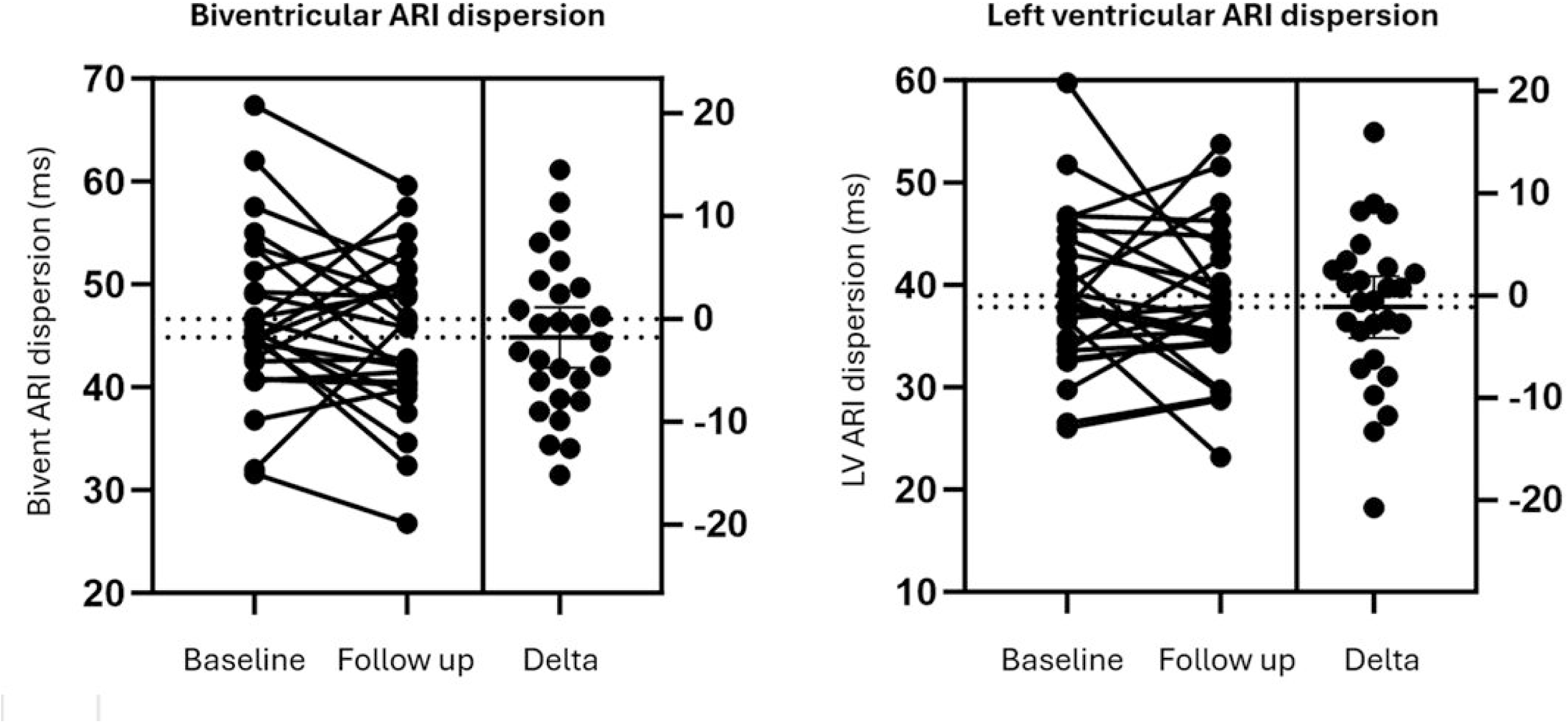
Correlation between delta LV ARI dispersion and delta ischemic burden, colour coded by above and below median scar.

### Predictors of improvement

At follow up, 15 patients had a reduction in LV ARI dispersion. These patients had lower baseline LVEF (26 vs 32%, p=0.02) and higher LVESVi (109 vs 78 ml/m^2^, p<0.01) compared to patients who failed to improve (Table 3). On variable importance analysis, baseline variables associated with improvement included LVESVi, LVEDVi, ischemia, scar and LVEF (Figure 5). At follow up, only change in ischemic burden and change in LVESVi were associated with improvement.

**Table 3.** Univariate predictors for response.

|  | Improved<br>(n=15) | Failed to improve<br>(n=12) | p value | Odds ratio |
| --- | --- | --- | --- | --- |
| Baseline factors |  |  |  |  |
| Age | 67.0 ± 9.8 | 66.5 ± 10.2 | 0.91 | 1.00 (0.92-1.07) |
| BMI | 27.5 ± 3.5 | 29.1 ± 4.6 | 0.32 | 0.91 (0.74-1.10) |
| Baseline LVEF | 25.9 ± 6.3 | 32.3 ± 6.7 | **0.02** | **0.87 (0.77-0.99)** |
| ACS presentation | 8 (57) | 3 (23) | 0.07 | 3.43 (0.66-17.9) |
| LVESVi | 109.0 ± 29.8 | 78.3 ± 22.9 | **<0.01** | **1.04 (1.01-1.08)** |
| Scar burden | 18.2 ± 12.3 | 13.7 ± 8.8 | 0.31 | 1.04 (0.97-1.12) |
| Ischemic burden | 57.1 ± 16.7 | 42.1 ± 20.5 | 0.06 | 1.04 (0.99-1.10) |
| Revascularization factors |  |  |  |  |
| Underwent CABG | 8 (53) | 3 (25) | 0.14 | 0.36 (0.07-1.82) |
| Revascularization index | 76.6 ± 17.1 | 63.8 ± 20.1 | 0.10 | 1.04 (0.99-1.09) |
| Delta LVESVi | -25.2 ± 17.5 | -7.9 ± 15.1 | **0.02** | **0.94 (0.89-0.99)** |
| Delta LVEDVi | -22.4 ± 21.5 | -2.7 ± 13.8 | **0.02** | **0.95 (0.90-1.00)** |
| Change in ischemic burden | -44.7 ± 20.5 | -22.5 ± 19.0 | **0.02** | **0.95 (0.90-1.00)** |

*Only 27 patients included as two did not have analysable paired ECGi data.*
*CABG: Coronary Artery Bypass Grafting, LVESVi: Indexed Left Ventricular End Systolic Volume, LVEDVi: Indexed Left Ventricular End Diastolic Volume*

**Figure 5.**
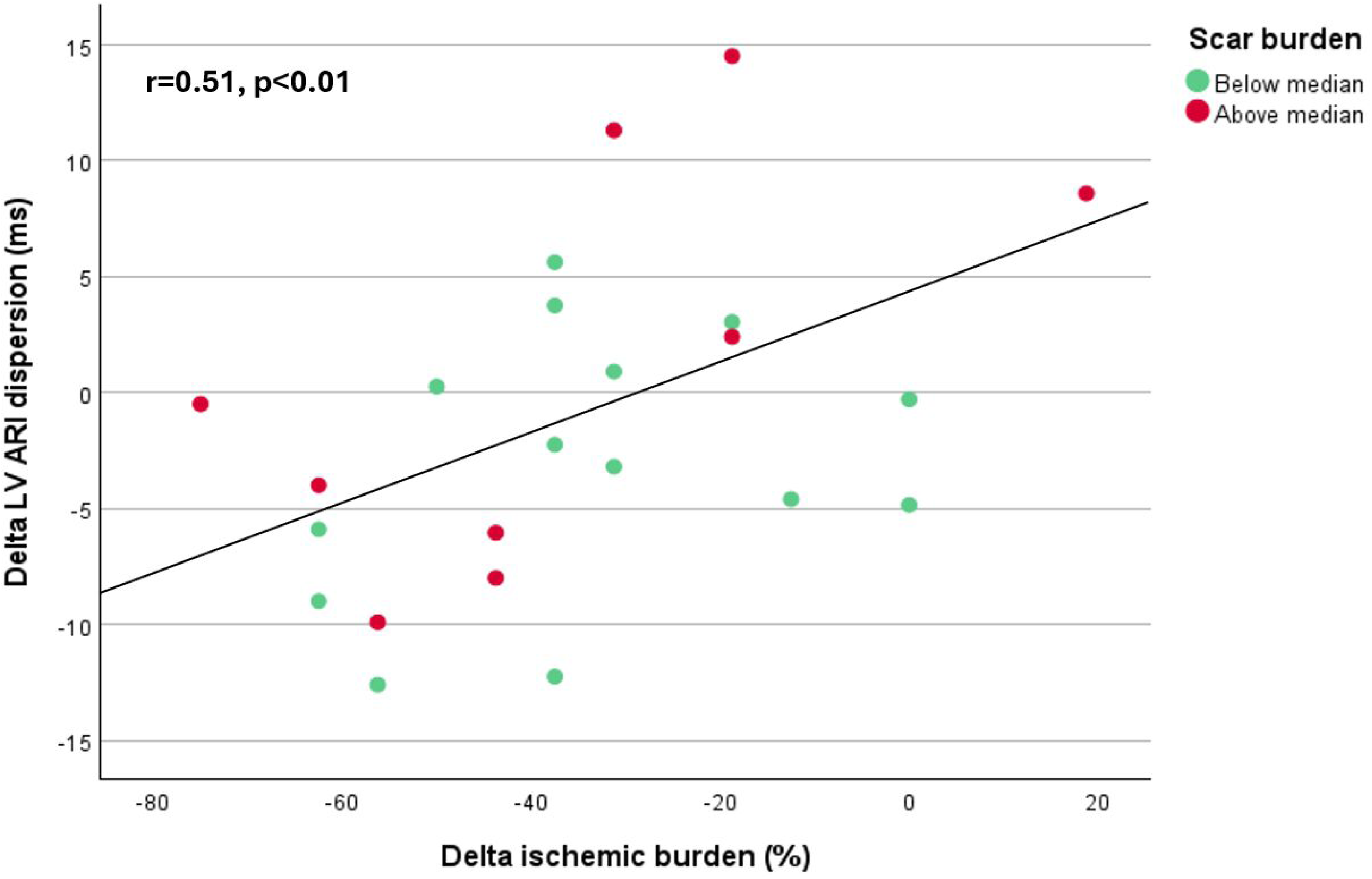
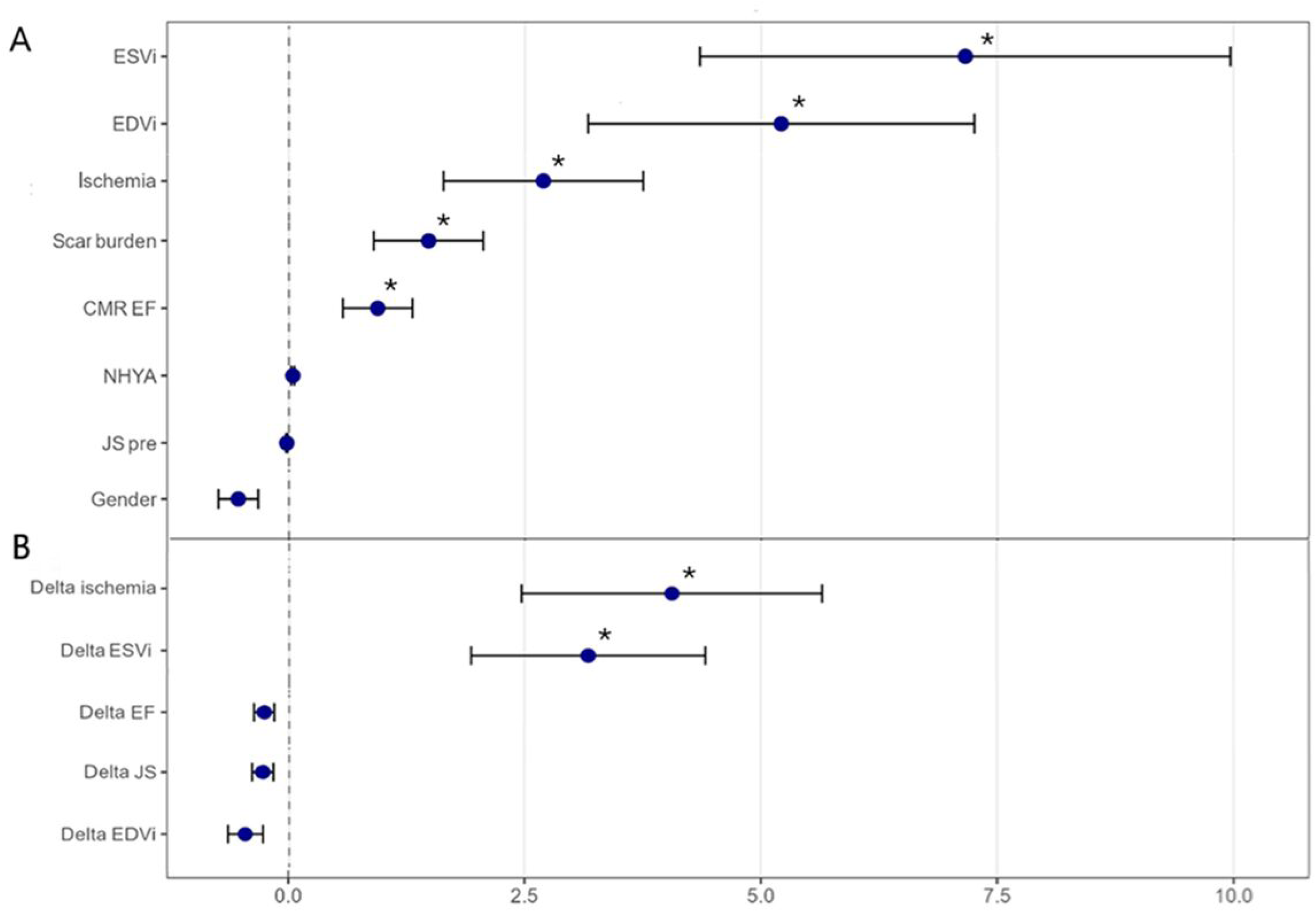
Random forest plots showing variable importance for response. A: Baseline variables, B: Delta variables.

On multivariate analysis, a model containing baseline ESVi, EDVi, ischemia and scar was highly predictive of improvement (AUC 0.92, CI 0.80-1.00, p<0.001)(Table S6).

## Discussion

This prospective analysis identified that revascularization was associated with reductions in LV volume and ischemia burden, as well as improvement in LVEF. However, it was not associated with a reduction in arrhythmic substrate across the whole cohort. However, delta arrhythmic substrate was heterogeneous, and factors associated with response included baseline and delta left ventricular volumes and ischemic burden.

### Baseline metrics

Scar was associated with the arrhythmic substrate at baseline, in keeping with the growing body of evidence that scar is highly predictive of arrhythmic events.^18^ Our work is consistent with other ECGi based studies that have identified increased ARI dispersion in patients with scar,^19^ as well as in CRT non responders with adverse remodelling.^15^

In this cohort, LVESVi was most strongly linked to the arrhythmic substrate, above LVEF. Similar findings have been reported previously.^20^ However, this was a cohort of exclusively severely impaired LVEF with a relatively narrow range; this may explain why it was no longer a predictive factor.

### Changes post revascularization

Improvements seen in LV function and size were greater than reported in other revascularization in ILVD studies, with STICH reporting an increase at 4 months of 2% and REVIVED a 4% increase at 6 months.^7,21^ Our study was made up of patients with recent acute coronary syndrome as well as stable patients; LV dysfunction in the former may have been due a combination of post infarct stunning and pre-existing systolic dysfunction, which may explain why these patients had larger reductions in LV volumes, increasing overall mean changes.^22,23^ It is also worth noting that the results of the main REVIVED trial were published whilst recruitment to this study was still ongoing. In a post REVIVED era, patients with ILVD were less likely to be considered for revascularization if they were stable and free of significant angina. This likely impacted the cohort being recruited to this study, supported by the high levels of angina at enrolment.

### Alterations in arrhythmic substrate

The identified association between change in ischemia and change in the arrhythmic substrate supports the underlying theory of ischemia-induced arrhythmogenesis, which outlines that ischemia depresses excitability of the myocardium, slows electrical conduction and increases heterogeneity of repolarisation.^24,25^

Delta LV ARI dispersion was chosen as the primary outcome as it has been linked to an increased risk of ventricular arrhythmias in patients with ILVD in previous invasive and non-invasive studies.^26^ Early ECG studies studied QT dispersion, identifying reductions after surgical revascularization ^27^, elective single vessel PCI ^28^ as well as chronic total occlusion PCI.^29^ Elliott et al identified that delta LV ARI dispersion on ECGi was associated with response to CRT and change in LVESVi, with non-responders demonstrating an increased dispersion at 6 months.^15^ However, delta findings have not been previously been reported on ECGi or invasive electrophysiology (EP) studies in relation to revascularization. There is therefore minimal published work regarding the test retest accuracy of such measures, however with development of reusable vests such literature is increasing.^30^

There are a number of explanations for why we did not see a reduction in arrhythmic substrate across the whole cohort, despite all patients achieving a high revascularisation index. Firstly, it is notable that almost a third of the cohort had a residual ischemic burden of ≥20% at follow up; this was similar regardless of revascularization modality. Very few trials have assessed delta ischemia burden and outcomes, most focusing on baseline ischemia only.^31^ The COURAGE nuclear study conducted myocardial perfusion imaging at baseline and post revascularization. At 12 months follow up, 16% of the PCI arm had >10% residual ischemia; the authors identified a survival benefit was only seen in those with ischemia reduction, this was amplified in those with significant demonstrable ischemia at baseline.^32^ In a study of patients with multi-vessel disease undergoing either PCI or CABG, delta ischemia burden on CMR was compared to an angiographic assessed revascularization index.^33^ Despite angiographic assessment judging 63% of patients had achieved complete revascularization, this was just 39% when assessed via CMR, with 34% having a residual ischemic burden greater than 33%.^33^ Residual microvascular disease is also likely to be a factor, how this impacts arrhythmic risk remains unknown.^34^

Notably, these cohorts included patients with preserved LV function; currently no data exists on the change in ischemic burden in the ILVD population undergoing revascularization; it is hoped some data will be provided by the ongoing STICH-3 trials.^35^

Secondly, scar is likely the main driver of arrhythmogenesis in this population. It may be that significant changes in dispersion were not seen due to the high burden of scar, which is not reduced by revascularization, on the contrary it can cause periprocedural infarction and new scar formation in some individuals, which could offset the benefits of reducing ischemic burden (Figure S3). Previous work has suggested that revascularization may also increase heterogeneity by creating pockets of viable tissue within scar.^36–38^

Baseline and delta LV volumes as well as ischemia burden were significant predictors of improvement in arrhythmic substrate. These factors may therefore be useful in identification of patients who may benefit from revascularization, however exact thresholds for these values remain unknown. Within our cohort, it was not until ischemic burden had reduced by more than 40% that reductions in arrhythmic substrate started to be seen (Central Illustration). Further work is needed to enable clinicians to prospectively identify patients who are likely to have a significant reduction in LV volumes and ischemic burden, who may then confer benefit in reduced arrhythmic risk.

### Study limitations

This study includes a small cohort of patients, and was only powered for identifying the correlation between delta LV ARI dispersion and delta ischemia.

Due to the study design, patients with a cardiac device in situ could not be recruited due to CMR compatibility requirements and therefore none of the patients within this cohort had a history of significant ventricular arrhythmia. Our findings therefore may not be applicable to these populations.

The measures of arrhythmic substrate used in this study, namely LV ARI dispersion, are surrogate markers of arrhythmic risk. Whilst there is strong evidence validating ARI dispersion with invasive EP metrics in cohorts with and without a history of arrhythmia, there is limited evidence linking ECGi metrics with long term clinical outcomes.

There is a risk of test retest variability in any repeated metric; to minimise this we ensured the same jacket size was used for both ECGi studies and utilised a standardised application protocol. We did not enrol healthy or control subjects, therefore we cannot confirm with certainty which of our findings are unique to the ILVD population. Given the limited range in LVEF within this cohort, it would be of interest to investigate patients with mild or moderately reduced LVEF to determine if there are differences in ECGi metrics between these patient groups, and additionally whether assessment of these metrics would enable us to re-stratify patients in the ‘grey zone’.

### Conclusions

The change in arrhythmic substrate, measured by LV ARI dispersion, was associated with the change in ischemic burden and change in LV volumes, however no significant decrease was noted in the cohort as a whole. There were significant rates of residual ischemia regardless of revascularization modality.

LV ARI dispersion is highly correlated with scar burden, which was not altered by revascularization. Further work is needed to prospectively identify patients likely to have a significant reduction in LV volumes and ischemic burden, who may confer benefit from coronary revascularization.

## Data Availability

Data will be made available upon reasonable request.

## Notes

### Competing Interest Statement

The authors have declared no competing interest.

### Clinical Trial

Not a trial

### Funding Statement

This work was funded by the British Heart Foundation (Clinical Research Training Fellowship FS 18/16/33396 and King?s BHF Center of Research Excellence Award RE/18/2/34213).

### Author Declarations

The study protocol was approved by the UK Health Research Authority in 2021; research ethics committee approval was gained prior to study commencement (REC reference: 21/SC/0024; IRAS 281825)

